# Benchmarking Deep Learning Models and Automated Model Design for COVID-19 Detection with Chest CT Scans

**DOI:** 10.1101/2020.06.08.20125963

**Authors:** Xin He, Shihao Wang, Shaohuai Shi, Xiaowen Chu, Jiangping Tang, Xin Liu, Chenggang Yan, Jiyong Zhang, Guiguang Ding

**Affiliations:** Department of Computer Science, Hong Kong Baptist University, Hong Kong, China; School of Automation, Hangzhou Dianzi University, Hang Zhou, China; School of Software, Tsinghua University, Beijing, China

**Author notes:** Corresponding author at Hong Kong Baptist University. Corresponding author at Hangzhou Dianzi University.

**Keywords:** COVID-19, Benchmark, Deep learning, Au-toML, Chest CT, Data augmentation

## Abstract

COVID-19 pandemic has spread all over the world for months. As its transmissibility and high pathogenicity seriously threaten people’s lives, the accurate and fast detection of the COVID-19 infection is crucial. Although many recent studies have shown that deep learning based solutions can help detect COVID-19 based on chest CT scans, there lacks a consistent and systematic comparison and evaluation on these techniques. In this paper, we first build a clean and segmented CT dataset called Clean-CC-CCII by fixing the errors and removing some noises in a large CT scan dataset CC-CCII with three classes: novel coronavirus pneumonia (NCP), common pneumonia (CP), and normal controls (Normal). After cleaning, our dataset consists of a total of 340,190 slices of 3,993 scans from 2,698 patients. Then we benchmark and compare the performance of a series of state-of-the-art (SOTA) 3D and 2D convolutional neural networks (CNNs). The results show that 3D CNNs outperform 2D CNNs in general. With extensive effort of hyperparameter tuning, we find that the 3D CNN model DenseNet3D121 achieves the highest accuracy of 88.63% (F1-score is 88.14% and AUC is 0.940), and another 3D CNN model ResNet3D34 achieves the best AUC of 0.959 (accuracy is 87.83% and F1-score is 86.04%). We further demonstrate that the mixup data augmentation technique can largely improve the model performance. At last, we design an automated deep learning methodology to generate a lightweight deep learning model MNas3DNet41 that achieves an accuracy of 87.14%, F1-score of 87.25%, and AUC of 0.957, which are on par with the best models made by AI experts. The automated deep learning design is a promising methodology that can help health-care professionals develop effective deep learning models using their private data sets. Our Clean-CC-CCII dataset and source code are available at: https://github.com/HKBU-HPML/HKBU_HPML_COVID-19.

## I. Introduction

The COVID-19 (Corona Virus Disease 2019) pandemic is an ongoing pandemic caused by severe acute respiratory syndrome coronavirus 2 (SARS-CoV-2) [1]. The SARS-CoV-2 virus can be easily spread among people via small droplets produced by coughing, sneezing, and talking [2]. Even worse, SARS-CoV-2 can be highly stable in a favourable environment so that it can adhere to different object surfaces up to several days [3], which causes a higher risk of getting infected by touching these contaminated surfaces and then touching their own faces.

COVID-19 is not only easily contagious, but also a serious threat to human lives. The COVID-19 infected patients usually present with pneumonia-like symptoms (fever, dry cough, dyspnea, etc.) and gastrointestinal symptoms such as diarrhea, followed by a severe acute respiratory infection. In some cases, acute respiratory distress accompanied by severe respiratory complications may even lead to death. According to the COVID-19 situation report [4] provided by the World Health Organization (WHO), as of the end of May, there were 5,934,936 COVID-19 infections and 367,166 deaths globally. The usual incubation period of COVID-19 ranges from one to 14 days. Many COVID-19 patients do not even know that they have been infected without any symptoms, which would easily cause delayed treatments and lead to a sudden exacerbation of the condition. Therefore, a fast and accurate method of diagnosing COVID-19 infection is crucial.

Currently, there are two commonly used methods for COVID-19 diagnosis. One is viral testing, which uses real-time reverse transcription-polymerase chain reaction (rRT-PCR) to detect viral RNA fragments. The other one is making diagnoses based on characteristic imaging features on chest X-rays or computed tomography (CT) scan images. [5] conducted the effectiveness comparison between the two diagnosis methods and concluded that chest CT has a faster detection from the initial negative to positive than rRT-PCR. However, the manual process of analyzing and diagnosing based on CT images highly relies on professional knowledge and is time-consuming to analyze the features on the CT images. Therefore, many recent studies have tried to use deep learning (DL) methods to assist COVID-19 diagnosis with chest X-rays or CT scan images.

However, the reported accuracy of the existing DL-based COVID-19 detection solutions spans a broad spectrum because they were evaluated on different datasets, making it difficult to achieve a fair comparison. In this paper, we aim to conduct a reproducible comparative study of DL methods for COVID-19 detection using chest CT scans. To this end, we first build a clean and segmented CT scans dataset based on a large-scale open-source dataset^1^ from CC-CCII (China Consortium of Chest CT Image Investigation) [6]. Our dataset, named Clean-CC-CCI, consists of three classes: novel coronavirus pneumonia (NCP), common pneumonia (CP), and normal controls (Normal). Totally, there are 340,190 slices of 3,993 scans from 2,698 patients in our dataset, where the number of slices of NCP, CP, and Normal is 131,517, 135,038, and 73,635, respectively. We split the dataset into the training and test sets according to the patient’s ID with a ratio of 4:1, the details of which are shown in Table II. Notice that our test set size is the largest one (e.g., it is twice of that in [6]), making our evaluation results more conservative than existing ones. Our benchmark dataset is made open to the public and can facilitate the fair comparison of new DL models for COVID-19 detection.

In this paper, we use our dataset to benchmark two types of state-of-the-art (SOTA) DL models: 1) 3D convolutional neural networks (CNNs), including DenseNet3D121 [17], R2Plus1D [18], MC3 18 [18], ResNeXt3D101 [17], Pre-Act ResNet [17], and ResNet3D series [17]; 2) 2D CNNs, including DenseNet121 [19], DenseNet201 [19], ResNet50 [20], ResNet101 [20] and ResNeXt101 [21]. We explore three key factors that may affect the detection performance, including model depth, methods of reading slice images, and model architecture. First, regarding the model depth, we compare the performance of the ResNet architecture [20] with 3D from 10 layers to 152 layers, i.e., ResNet3D10, ResNet3D18, ResNet3D34, ResNet3D50, ResNet3D101, ResNet3D152, and ResNet3D200. Second, in terms of how to read the slice images, we consider two popular approaches: one is to read a slice as an RGB image with three channels; another is to convert the slice to a greyscale image with only one channel. Therefore, the scan images used to train the model will be different because of the different ways of slice reading. Third, we exploit multiple DNN architectures including the hand-craft models and automatically generated models with AutoML techniques [22], [23]. We use seven 3D models to analyze the effect of two types of scan data. Besides, we discuss the influence of the number of slices in a CT scan on the model performance. We also evaluate the effectiveness of the mixup data augmentation method by comparing model accuracy before and after applying the mixup method. Our major contributions are summarized as follows:

1. We build an open benchmark dataset Clean-CC-CCII for COVID-19 detection using chest CT scans, and benchmark 9 different CNN architectures with more than 20 variants.
2. We find that both 3D and 2D CNNs are promising solutions for detecting COVID-19 infections. However, the overall performance of 3D CNNs is better than 2D CNNs. Besides, the results of the ResNet3D series show that the model performance does not scale very well with the model depth.
3. We find that the models can achieve higher AUC when the slices are converted to greyscale images.
4. To the best of our knowledge, this is the first paper to explore the relationship between model performance and the number of slices in a CT scan. Our result shows that there is no significant correlation between them. In other words, increasing the number of slices does not necessarily improve the model performance. Instead, the model trained on scan data with a small number of slices can also achieve comparable or even better results.
5. We demonstrate that the mixup data augmentation method [24] can effectively improve model accuracy in our study.
6. We develop an automated deep learning methodology to generate a lightweight deep learning model MNas3DNet41. On our dataset, it achieves an accuracy of 87.14%, F1-score of 87.25%, and AUC of 0.957, which are on par with the best results of the highly fine-tuned models made by AI experts.

The rest of the paper is organized as follows. Section II describes the related work. In section III, we describe the strategies used to build our dataset, the comparison study of SOTA CNN models, and the automated model design methodology. Section IV presents and discusses the experimental results. We conclude the paper and introduce the future research directions in Section V.

## II. Related work

In recent years, DL techniques have been proved to be effective in the diagnosis of diseases with X-ray and CT images [25]. To enable machine learning techniques be applied in helping detect COVID-19, an increasing number of publicly available COVID-19 datasets has been proposed in the past few months as shown in Table I. These datasets can be classified into two classes: X-ray and CT scan images. Machine/Deep learning techniques highly rely on both the quality and quantity of the dataset.

**TABLE I.** Summary of the existing studies of deep learning-based methods for COVID-19 detection. ‡: the number of scans. *: the number of patients.

| Paper | Type | 2D/3D dataset | Download-able? | Dataset Statistics |  |  | Class Statistics (#slices) |  |  | Size of Test Set | 2D/3D model? | Accuracy (%) | AUC |
| --- | --- | --- | --- | --- | --- | --- | --- | --- | --- | --- | --- | --- | --- |
|  |  |  |  | #patients | #scans | #slices | NCP | Non-NCP |  |  |  |  |  |
|  |  |  |  |  |  |  |  | CP | Normal |  |  |  |  |
| [7] | X-ray | 2D | Yes | - | - | 1,531 | 100 | 1,431 |  | 764 | 2D | - | 0.9172 |
| [8] | X-ray | 2D | Yes | - | - | 5,941 | 68 | 4,290 | 1,583 | 1,188 | 2D | 88.39 | - |
| [9] | X-ray | 2D | Yes | - | - | 100 | 50 | 50 |  | 20 | 2D | 98.00 | - |
| [10] | CT | 2D | No | - | - | 133 | 68 | 65 |  | 26 | 2D | 93.20 | - |
| [11] | CT | 2D | No | 194 | - | 1,020 | 510 | 510 |  | 102 | 2D | 99.63 | 0.994 |
| [12] | CT | 2D | Yes | - | - | 425 | 178 | 247 |  | 45 | 2D | 98.78 | - |
| [13] | CT | 2D | Yes | 143 | - | 746 | 349 | 397 |  | 186 | 2D | 86.00 | 0.91 |
| [14] | CT | 2D | Yes | - | - | 746 | 349 | 397 |  | 105 | 2D | 87.60 | 0.961 |
| [15] | CT | 3D | No | 542 | 630 | - | 313* | 229* |  | 131* | 3D | 90.10 | 0.975 |
| [16] | CT | 3D | No | 3,322 | 4,356 | - | 1,296‡ | 1,735‡ | 1,325‡ | 427‡ | 3D | - | 0.96 |
| [6] | CT | 3D | Yes | 2,778 | 4,356 | 444,034 | 1,578‡ | 1,614‡ | 1,164‡ | 389‡ | 3D | 92.49 | 0.976 |
| Ours | CT | 3D | Yes | 2,698 | 3,993 | 340,190 | 1,515‡ | 1,513‡ | 965‡ | 798‡ | 2D&3D | 88.63 | 0.959 |

### A. Publicly-available datasets of COVID-19

IEEE8023 Coivd-chestxray-dataset [26] is an open dataset of COVID-19 cases with chest X-ray and CT images, which allows users to submit other COVID-19 data to this dataset. However, this dataset mainly focuses on X-ray images with only a very small number of CT scans. Based on this dataset, several DL based techniques have been proposed [7]–[9] to detect COVID-19.

Covid-ct-dataset [27] is a CT dataset of COVID-19, which is mainly composed of CT images extracted from PDF files of COVID-19 papers in medRxiv and bioRxiv. Thus, it has two main drawbacks. First, many CT images contain some marks created by the CT machine or doctors, which may have a high impact on the DL techniques. Second, each patient has only one to several CT images instead of a complete 3D scan volume, which results in some difficulties to use 3D CNNs to exploit the depth information of the lung.

CC-CCII is another publicly available CT volume dataset proposed by [6]. It is currently one of the largest CT datasets for COVID-19, which contains 617,775 slices of CT images from 6,752 scans of 4,154 patients. It has 3 classes of novel coronavirus pneumonia (NCP), common pneumonia (CP), and normal controls (Normal). CP includes bacterial pneumonia and viral pneumonia. However, this dataset (version 1.0 released on 23 April 2020) contains some errors (e.g., disorder of CT images in some scans, some scans include CT of the head but not the lung, etc.).

COVID-19-CT-Seg-Dataset [28] is a publicly available CT dataset of COVID-19. It contains 20 well-labeled scans with annotation of left lung, right lung and lesions. Three experienced radiologists are involved for each annotation: two radiologists do the annotation and one does the verification.

### B. DL-based methods for COVID-19 detection

Most research is conducted on CT images, but many of them do not exploit the 3D information of CT images, such as the work by [10], [13], [14]. They only propose the DL models with 2D CNNs for COVID-19 detection. [11] is the most related work to ours; but it only benchmarks ten 2D CNNs and compares their performance in classifying 2D CT images on their private dataset with 102 testing images.

On the other hand, the studies in utilizing 3D CT images are relatively rare, which is mainly due to the lack of 3D CT scan dataset of COVID-19 in the earlier days. However, there still exist some work proposing 3D CNNs with their private 3D CT datasets (e.g., [16], [15]). Recently, [6] publish a large-scale publicly available 3D CT dataset, based on which they propose 3D CNNs methods to segment lesion and detect COVID-19. However, in [6], only two DL models are exploited to evaluate the model performance using 10% of the dataset as test set. It is of practical importance to evaluate which types of models are suitable to the 3D CT images in detecting COVID-19.

There are also some other studies conducted on X-ray images. For example, [9] propose three 2D CNNs for COVID-19 detection. [7] introduce a deep anomaly detection model for fast and reliable screening. [8] investigate the estimation of uncertainty and interpretability by droiopweights-based Baysian CNN on the X-ray images. [12] use both X-ray images and CT images to do segmentation and detection.

### C. Automated model design for medical image analysis

In recent years, Automated Machine Learning (AutoML) has created many SOTA results by automatically searching model architectures and hyper-parameters for specific tasks [22], [23], [29]. For example, [30] introduce AutoML into the medical image processing task. They used five public datasets, MESSIDOR, OCT images, HAM 10000, Paediatric images and CXR images, to train models by Google Cloud AutoML. Their experimental results demonstrate that AutoML can generate competitive classifiers compared to manually designed DL models.

## III. Materials and methods

### A. Dataset

[6] provide an open-source chest CT image dataset for COVID-19 diagnosis, namely China Consortium of Chest CT Image Investigation (CC-CCII), which contains a total of 617,775 CT slices of 6,752 CT scans from 4,154 patients. CC-CCII has three classes: novel coronavirus pneumonia (NCP), common pneumonia (CP), and normal controls (Normal). CP includes bacterial pneumonia and viral pneumonia. To best of our knowledge, CC-CCII is the largest COVID-19 CT dataset which is publicly available currently. It would be helpful for accelerating the research on machine learning based methods in COVID-19 diagnosis. However, CC-CCII has five main issues (i.e., damaged data, non-unified data type, repeated and noisy slices, disordered slices, and non-segmented slices) that would have high negative impacts on the model performance. In this section, we first describe our methods to address the problems in CC-CCII to generate a better dataset for DL techniques. Then we introduce the strategies of scan images construction.

#### 1) Data pre-processing

##### Damaged data

According to the work in [6], the authors claim that the original number of CT slices in NCP, CP, and Normal is 164,241, 183,933, and 95,860, respectively. However, the dataset provided by [6] is composed of many zip files, some of which are damaged (i.e., around 10% of downloaded files cannot be successfully decompressed). As a result, we only obtain 397,945 CT slices, where there are 152,791, 157,940, and 87,214 slices in the class NCP, Normal, and CP, respectively.

##### Non-unified data type

In CC-CCII, there are various types of image files, such as “tif”, “jpg”, “bmp”, and “png”. For the same type of image files, some suffix names are also different, e.g., “0001.jpg” and “0002.JPG”, which is not friendly to reading data. Therefore, we unify all slice image files to the PNG format with the suffix “.png” without losing information of the original files.

##### Repeated & noisy slices

The third problem is that there exist duplicated slices in many CT scans. For example, in the class of Normal of CC-CCII, the scan ID of 199 in the patient ID of 764 has the repeated images (file names: 68-135) that are the same with file names: 1-67. For the DL-based methods using 3D CT volumes, the repeated slices are very noisy to the models since the 3D CT volume with repeated slices may form two lungs. Some repeated scans also affect the shape of the lung in the CT volume so that the model could easily fail to correctly extract the features. Thus, we manually remove those duplicated slices to avoid redundant data by scanning all the images in CC-CCII. In addition, some CT images of CC-CCII contain unrelated body parts (e.g., the head or buttocks) that are not related to the lung. For example, in the class of CP, scan 3098 in patient 736 has the head part in CT images “0000.png” and “0011.png”.

##### Disordered slices

In CC-CCII, the slices should have been organized in the same order for different scans so that the software can read the images according to the file name to generate a complete 3D lung. For example, by default, “0001.png” is regarded as the first slice followed by “0002.png, 0003.png, …”. However, the images in some scans are disordered so that these images cannot be constructed as the same shape of 3D lung with other scans with correct orders. These disordered slices could give confusing information to the DL models. Therefore, we go through all scans of CC-CCII and rearrange the disordered slices to the correct order.

##### Lung segmentation

The last problem is that in some slices, the lungs are segmented, while some are not. The segmented lungs and un-segmented lungs have very clear difference that the un-segmented lungs have clear white borders out of the lung regions while segmented lungs have not. In order to keep the consistency for all slices, we use an open-source K-Means based method ^2^ to segment lungs form the CT slices and remove the white background. Fig. 1 presents some examples of CT scan of class NCP, CP, and Normal after segmentation.

**Fig. 1.**
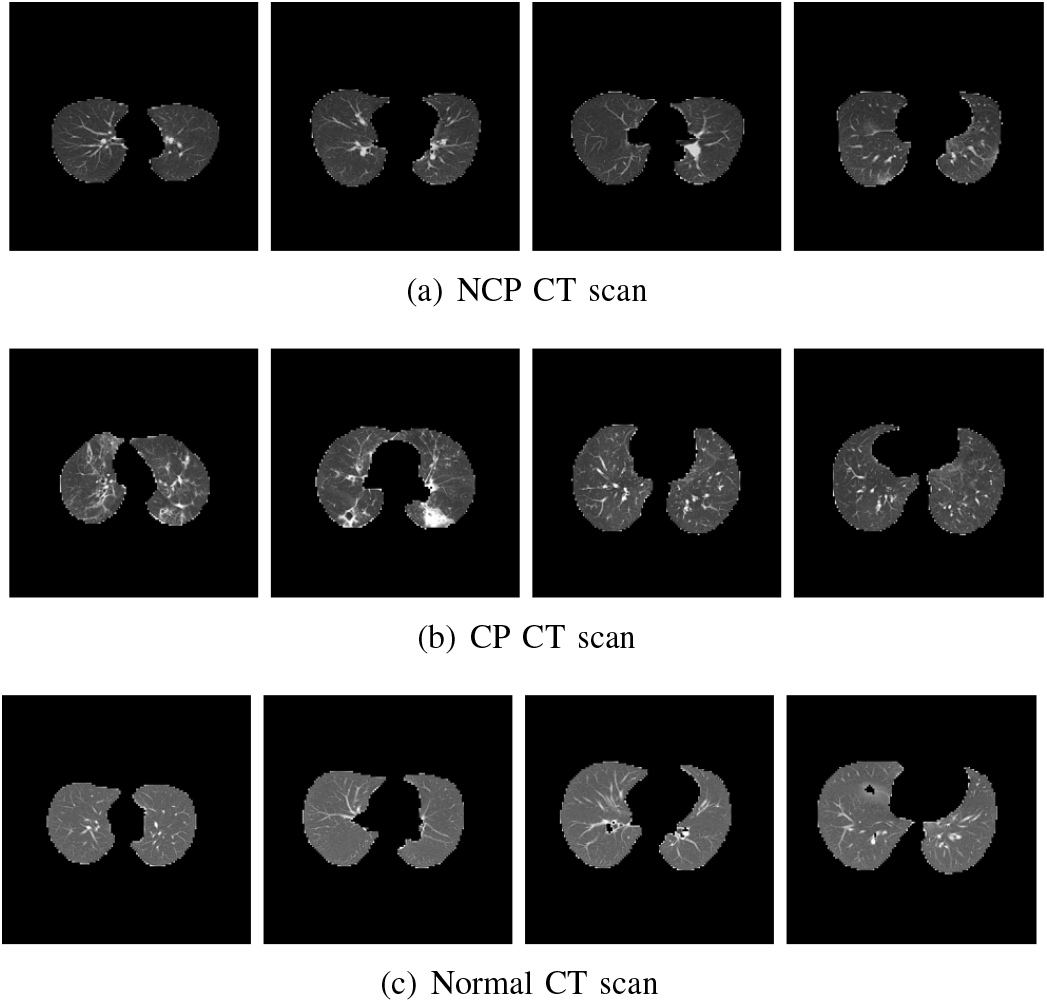
Some samples of segmented chest CT scan images of class (a) NCP, (b) CP, and (c) Normal.

After addressing the above problems, we construct a clean CC-CCII dataset named Clean-CC-CCII, which is more suitable to DL-based methods in COVID-19 diagnosis. The statistics of our dataset are presented in Table II. Finally, our Clean-CC-CCII dataset consists of 340,190 slices of 3,993 scans from 2,698 patients. The dataset is divided into the training set and the test set according to patients to make sure that the CT scan images from the same patient will appear either only in the training set or in the test set. The ratio of the number of scans in the training set and the test set is 4:1.

**TABLE II.** The statistics of our Clean-CC-CCII dataset. The dataset is divided into the training and test sets. The ratio of the number of patients, scans and slices for the two sets is 4:1. NCP, CP, AND Normal indicate novel coronaviruspneumonia, common pneumonia, and normal control, respectively.

| Classes | #Patients |  | #Scans |  | #Slices |  |
| --- | --- | --- | --- | --- | --- | --- |
|  | Train | Test | Train | Test | Train | Test |
| NCP | 726 | 190 | 1,213 | 302 | 106,075 | 25,442 |
| CP | 778 | 186 | 1,210 | 303 | 106,878 | 28,160 |
| Normal | 660 | 158 | 772 | 193 | 59,164 | 14,471 |
| Total | 2,164 | 534 | 3,195 | 798 | 272,117 | 68,073 |

#### 2) Scan images construction

After data pre-processing, we need to construct CT scan images as inputs of DL models for training. As shown in Fig. 3, there are two steps before feeding data into DL models: slice sampling and slice processing.

##### Slice sampling

In our dataset, each CT scan contains a different number of slices as shown in Fig. 2. The minimum and maximum number of slices are 9 and 457, respectively. However, DL models generally require the same dimensional inputs. To keep the same dimension inputs, we propose two types of slice sampling strategies: random sampling and symmetrical sampling. Specifically, the random sampling strategy is applied to the training set, which can be regarded as data augmentation, while the symmetrical sampling strategy is performed on the test set to avoid introducing randomness into the testing results. Besides, because the number of slices can be manually set to different values, both sampling strategies support automatically select upsampling or downsampling based on the original and target number of slices. We will also study the performance impact of the number of slices in Section IV-C. Notably, the relative order between slices remains the same before and after sampling. The details of our sampling strategies are given in Algorithms 1 and 2 of A.

**Fig. 2.**
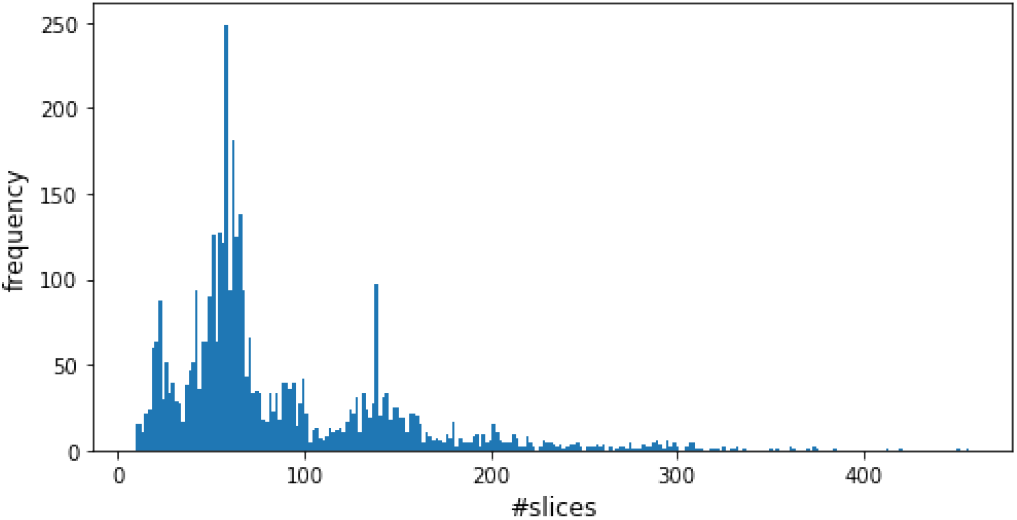
The frequency of the different number of slices. The minimum and maximum number of slices are 9 and 457, respectively.

##### Slice processing

After slice sampling, each scan data is composed of the same number of slices. We then resize all slices to 160 × 160 and central crop to 128 × 128. In this way, the final input data sizes for the 3D and 2D models are *c* × *d* × 128 × 128 and *d* × 128 × 128, respectively, where *c* ∈ {1, 3} is the number of channels of the slice image, and *d* indicates the configured number of slices. For all scan data in the training set, we apply a 3D random horizontal flip transformation. The scan data in both the training and test sets is normalized by subtracting the mean and dividing the variance.

### B. A comparative study of COVID-19 detection methods

In this study, we aim to investigate the performance of different types of DL models on detecting COVID-19 infection with chest CT scans. Therefore, we implement various experiments to evaluate the potential effective methods for COVID-19 diagnosis. Specifically, we compare the performance between SOTA DL models, including 3D and 2D models, and explore the relationship between model performance and (a) model depth and (b) how to read slice images. We also evaluate the effectiveness of the mixup data augmentation method in improving model classification accuracy.

Our pipeline of using DL models to classify CP, NCP, and Normal CT scans is shown in Fig. 3. The first step is to construct CT scan images to feed into the DL models by slice sampling and processing. The sizes of all slices are fixed to 128 × 128 for the model inputs. The models are trained with the training set and evaluated on the test set.

**Fig. 3.**
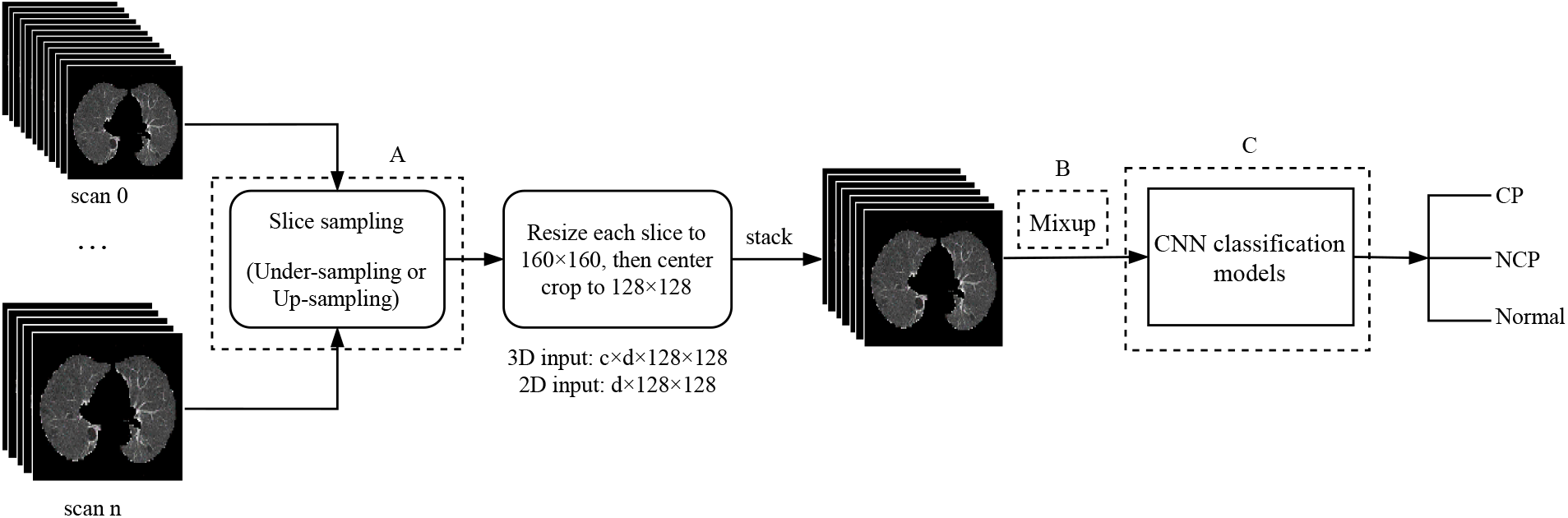
The pipeline of DL models to detect novel coronavirus pneumonia (NCP), common pneumonia (CP), and normal controls (Normal) from chest CT scans. For each step, the input data of the models is a batch of scan images. All scan images contain the same number of slices by sampling, and the size of each slice is fixed to 128 *×* 128. The final input data sizes for the 3D and 2D models are *c × d ×* 128 *×* 128 and *d ×* 128 *×* 128, respectively, where *c* is the number of channels of slice images, and *d* indicates the number of slices in a scan data. Three dashed boxes represent three different experiments: 1) the box A explores the effect of different sizes of scan data on model performance by sampling the different number of slices.; 2) the box B evaluates the effectiveness of mixup data augmentation method; 3) the box C compares the performance of different models and the performance of models at different depths.

#### 1) Exp 1: Comparing different CNN models

In this study, we evaluate 17 CNN classification models shown in Table III, including 3D models and 2D models. For the 3D models, we use DenseNet3D121 [17], R2Plus1D [18], MC3 18 [18], ResNeXt3D101 [17], PreAct ResNet3D [17], and ResNet3D series [17] (ResNet3D10, ResNet3D18, ResNet3D34, ResNet3D50, ResNet3D101, ResNet3D152, and ResNet3D200). For the 2D models, we use DenseNet121 [19], DenseNet201 [19], ResNet50 [20], ResNet101 [20] and ResNeXt101 [21].

**TABLE III.**
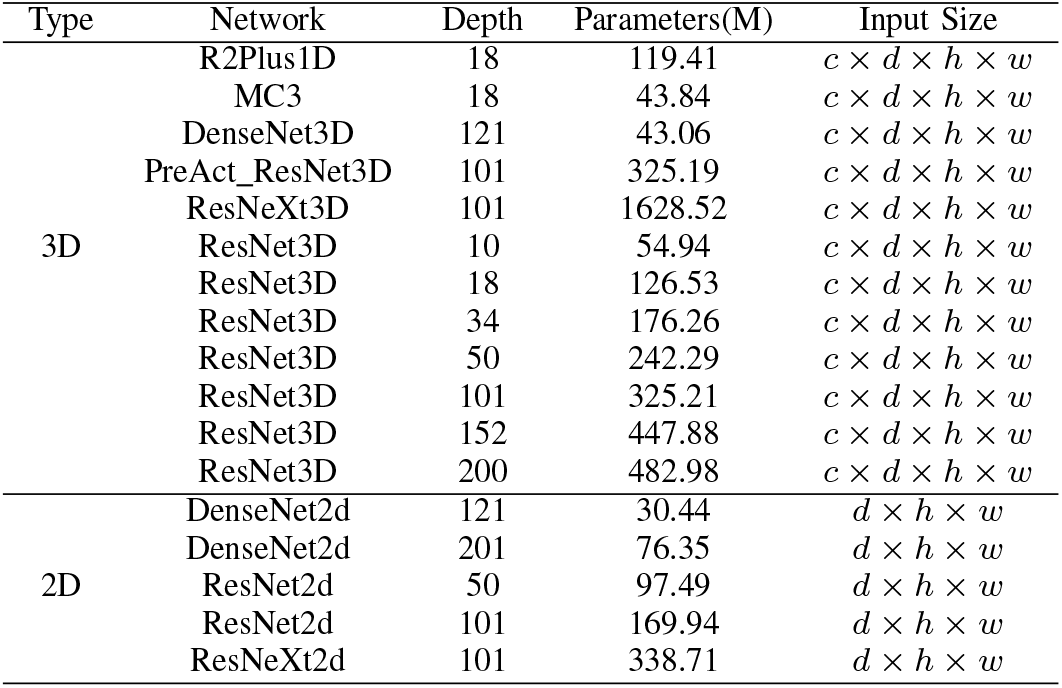
The CNN classification models used in our study. The input sizes for 3D and 2D models are *c × d × h × w* and *d × h × w*, respectively, where *c* is the number of channels of the slice image, *d* is the number of slices in a scan image, *h* and *w* indicate the height and width of the slice image, respectively.

For the 2D models, the input scan data is composed of greyscale slice images. In terms of 3D models, we evaluate two types of scan data: RGB slice images with three input channels and greyscale slice images. Besides, for both 2D and 3D models, the size of slices is fixed to 128 × 128. Therefore, the input sizes for 3D and 2D models are *c* × *d* × 128 × 128 and *d* × 128 × 128, respectively, where *c* is the number of channels of the slice image that depends on how to read the slice images, and *d* is the number of slices in a scan image. *c* = 3 and *c* = 1 indicate that each slice is read as the RGB and greyscale image, respectively. The number of input channels in the first convolutional layer of all models is modified accordingly to handle the input with different size.

#### 2) Exp 2: Comparing the different number of slices

In [6], the scan input is fixed to 64 slices. However, in our dataset, the number of slices contained in different CT scans ranges from 9 to 457, and the mean value is 85 as shown in Fig. 2. Intuitively, the higher number of slices, the more information can be extracted by the models, which could result in a higher performance. We empirically study the performance impacts of the number of slices by setting *d* to different values. We choose four representative 3D models (MC3 18, DenseNet3D121, ResNet3D101, and ResNeXt3D101) to evaluate the relationship between the model performance and the number of slices. For MC3 18, DenseNet3D121, and ResNet3D101, we evaluate five types of scan images containing 16, 32, 64, 128, and 256 slices, respectively. For ResNeXt3D101, it is too large to fit into the GPU memory when *d >* 64, so *d* is chosen with 16, 32, and 64.

#### 3) Exp 3: Training with mixup

Mixup [24] is a generic and straightforward data augmentation strategy, which has been proven to be effective in improving the model performance on 2D image classification tasks. Therefore, we explore the effectiveness of the mixup method in our 3D CT scan classification task.

In essence, mixup trains a DL model on linear combinations of pairs of examples and their labels. The formula is given as follows:

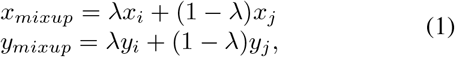

where (*x*_*i*_, *y*_*i*_) and (*x*_*j*_, *y*_*j*_) are two feature-target vectors drawn at random from the training set, and the variable *λ* ∈ [0, 1] obeys a *β*-distribution, i.e., *λ* ∼ *β*(*α, α*) for *α* ∈ (0, ∞). By doing so, a new feature-target vector will be generated by mixing up two feature-target vectors, which encourages the model to behave linearly in-between training examples.

In our experiments, we also use four representative 3D models (MC3_18, R2Plus1D, ResNet3D101, DenseNet121) to evaluate the feasibility of the mixup strategy. We set *α* = 0.4, as recommended in [24].

### C. Automated model design for COVID-19 detection

The results of all baseline experiments (to be discussed in Section IV) show that DL is a powerful tool to assist the detection of COVID-19 infection based on CT images, where 3D models generally outperform 2D models. However, as shown in Table III, 3D models have a very large model size and are slow to train. Based on the results of Table IV and Table V, we can see that a larger or a deeper model does not necessarily result in better performance. For example, ResNeXt3D101 is the largest model in our evaluated models, but its performance is not the best. Therefore, in this section, we aim to design a lightweight 3D model, which is expected to achieve comparable or even better results than the baseline 3D models and is easier to deployment for faster detection.

**TABLE IV.**
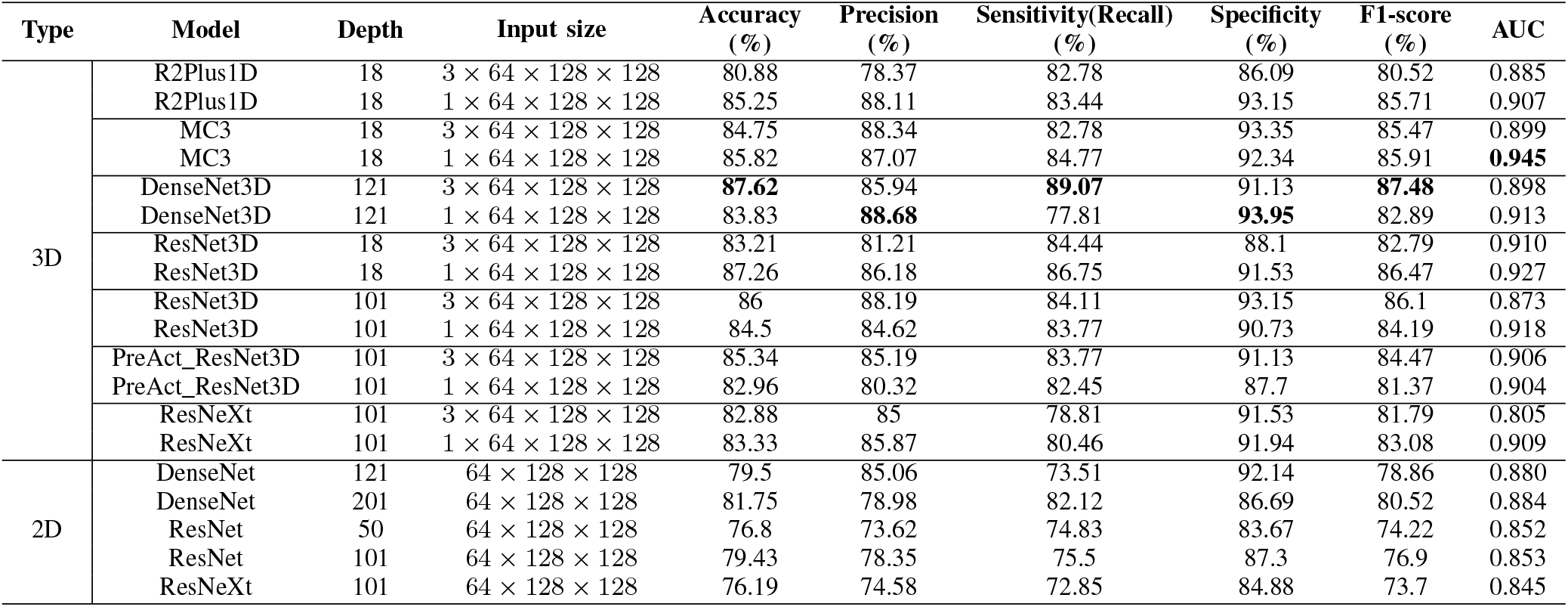
The performance comparison between different CNN classification models on the test set. The positive and negative cases are assigned to NCP and non-NCP (I.E., CP and Normal) scans, respectively. For 3D models, there two types of input size: 3 and 1 indicate that the scan data is composed by RGB and greyscale slice images, respectively.

**TABLE V.** The performance comparison between ResNet3D models of different depths on the test set. For all models, the input scan data has the same number of slices, i.e., 64. “Accuracy” indicates the final value of all data, while for metrics such as “precision, sensitivity, specificity, f1-score", NCP is the positive sample, and non-NCP (CP and Normal) is the negative sample.

| Model | Depth | Input size | Accuracy (%) | Precision (%) | Sensitivity(Recall) (%) | Specificity (%) | F1-score (%) | AUC |
| --- | --- | --- | --- | --- | --- | --- | --- | --- |
| ResNet3D | 10 | $1 \times 64 \times 128 \times 128$ | 85.35 | 79.68 | 83.11 | 87.10 | 81.36 | 0.918 |
| ResNet3D | 18 | $1 \times 64 \times 128 \times 128$ | 87.26 | 86.18 | **86.75** | 91.53 | 86.47 | 0.927 |
| ResNet3D | 34 | $1 \times 64 \times 128 \times 128$ | **87.83** | 85.34 | **86.75** | 90.93 | 86.04 | **0.959** |
| ResNet3D | 50 | $1 \times 64 \times 128 \times 128$ | 86.61 | 88.15 | 83.77 | **93.15** | 85.91 | 0.924 |
| ResNet3D | 101 | $1 \times 64 \times 128 \times 128$ | 84.50 | 84.62 | 83.77 | 90.73 | 84.19 | 0.918 |
| ResNet3D | 152 | $1 \times 64 \times 128 \times 128$ | 85.46 | 85.57 | 86.42 | 91.13 | 86.00 | 0.915 |
| ResNet3D | 200 | $1 \times 64 \times 128 \times 128$ | 87.19 | **88.70** | 85.76 | 93.35 | **87.21** | 0.927 |

However, manually designing a deep neural network is a time-consuming process that highly relies on experience and expertise. Luckily, a recent technique, namely neural architecture search (NAS), would be a promising solution for us. NAS can be seen as a sub-field of AutoML [22], [23], [29], which draws much attention from academia and industry as it can design various neural networks automatically. In the following content, we first introduce our search space and search strategy, and then describe the implementation details and experimental results.

#### 1) Search space

The first step of NAS is to build the search space, which defines the design principles of neural architectures. MobileNet [31] and MobileNetV2 [32] are a class of efficient models manually designed for mobile and embedded devices for efficient inference. Many NAS studies [33], [34] use the MobileNetV2 structure to design the factorized hierarchical search space, but they mainly focus on 2D image recognition tasks. In this work, we also exploit MobileNetV2 as the backbone to design the 3D search space.

An overview of the final model is shown in Fig. 4, which consists of *n* different cells. The number of blocks in a cell can be different, represented by [*B*_1_, …, *B*_*i*_, …, *B*_*n*_]. The stride is set to 2 in the first block if the resolutions of input and output are different, and the stride is 1 in all other blocks. The blocks within the same cell have the same number of input/output channels. Besides, the structure of each block is selected from a series of 3D mobile inverted bottleneck convolution operations [32], represented by *K* × *K_MBConvE*, where *K* is the filter kernel size and *E* is the expansion ratio of linear layers. In our method, the search space consists of the following operations: {3 × 3_*MConv*3, 3 × 3_*MConv*4, 3 × 3_*MConv*6, 5 × 5_*MConv*3, 5 × 5_*MConv*4, 7 × 7_*MConv*3, 7 × 7}_*MConv*4.

**Fig. 4.**
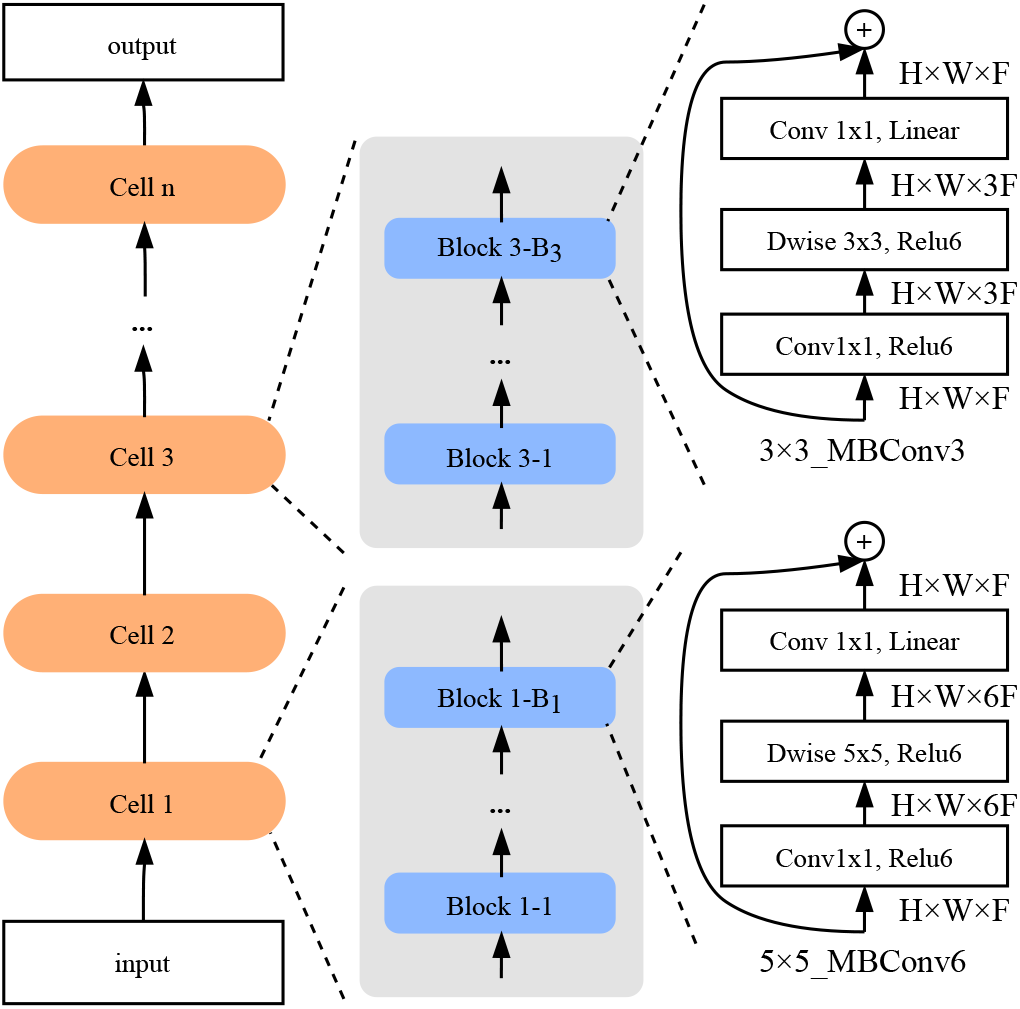
The overview of our search space. The model is generated by stacking a predefined number of cells, and each cell contains a different number of repeated identical blocks. Only the first block has a stride of 2 if resolutions of input and output are different, but all other blocks have a stride of 1.

#### 2) Search strategy

After building the search space, we can see that the key idea of the search task is to select the best submodel (in terms of validation accuracy) from the super-model. As summarized in [22], [29], there are various of search strategies, such as reinforcement learning, evolutionary algorithms, gradient descent-based methods, and random search. In recent studies [35], [36], the authors demonstrate that random search is a more competitive method than many others. Therefore, we also apply the random search strategy.

#### 3) Implementation details

The pipeline of our NAS methodology is shown in Fig. 5, which contains two stages for searching 3D models on our Clean-CC-CCII dataset: the search stage and the evaluation stage.

**Fig. 5.**
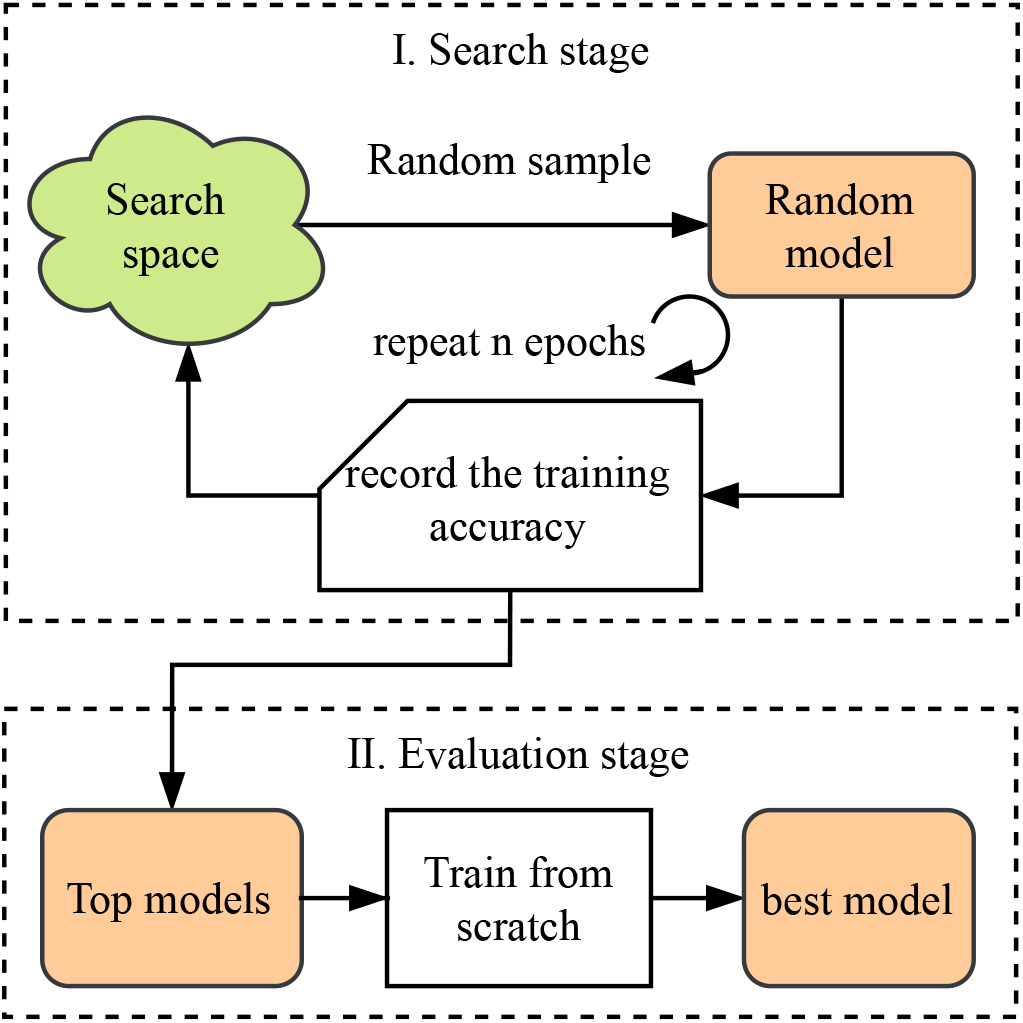
The pipeline of random strategy of searching for 3D models on our dataset. There are two stages: 1) search and 2) evaluation stage. The search stage is to select some promising neural networks, which are then evaluated in the evaluation stage.

### Search stage

In the search stage, we search for 100 epochs. Each epoch consists of a number of steps. We sample a new neural architecture every five steps and make sure that every sampled architecture is trained. Note that only the training set is used for training and evaluating the sampled models in the search stage. At the end of the search stage, there are 100 neural architectures and their corresponding training accuracy. **Evaluation stage**. After the search stage, we need to select several top ranked models (in terms of validation accuracy) for the next stage. Specifically, according to the training records, we choose those models that perform better validation accuracy than the previous sampled models. The selected models are first trained with the training set from scratch for 200 epochs, and then evaluated on the test set.

### Implementation details

For both search and evaluation stages, we use the Adam optimizer [37] with an initial learning rate of 0.001. The batch size is 64. The loss function is crossentropy. All input CT scan samples consist of 64 greyscale slice images. The number of cells is fixed to 6. We design two experiments to search for models with different depths: one is [4, 4, 4, 4, 4, 1] (21 layers), the other is [8, 8, 8, 8, 8, 1] (41 layers). For both cases, the number of channels of each block in different cells is [24, 40, 80, 96, 192, 320]. Each experiment is conducted on four Nvidia Tesla V100 GPUs (32-GB version). Furthermore, to improve the searching efficiency, we fix the height and width of the input scan to 60 × 60 during the search stage, and restore the size to 128 × 128 in the evaluation stage.

Our NAS-related code is based on NNI^3^ and can be found at https://github.com/arthursdays/HKBU_HPML_COVID-19.

## IV. Results and discussion

In this section, we present and analyze the results of the different experiments mentioned above. All models are trained using the Adam [37] optimizer with an initial learning rate of 0.001. The cosine annealing scheduler [38] is applied to adjust the learning rate.

### A. Evaluation metrics

To compare the performance of CNN models, we use several commonly used evaluation metrics as follows:

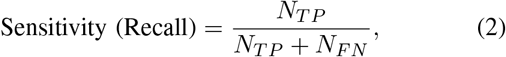

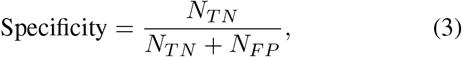

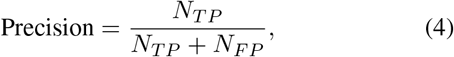

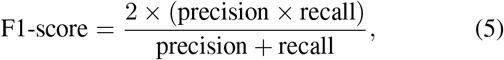

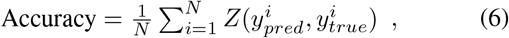

where 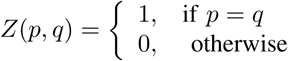.

Besides, the area under the receiver operating characteristic (ROC) curve (AUC) is also applied to evaluate the performance of COVID-19 diagnosis. In this study, the positive and negative cases are assigned to NCP and non-NCP (i.e., CP and Normal) scans, respectively. Specifically, *N*_*TP*_ and *N*_*TN*_ indicate the number of correctly classified NCP and non-NCP scans, respectively. *N*_*FP*_ and *N*_*FN*_ indicate the number of wrongly classified NCP and non-NCP scans, respectively. The accuracy is the micro-averaging value for all test data, which is used to evaluate the overall performance.

### B. Results of Exp 1: Comparing different CNN models

The performance comparison between different CNN modes including 3D and 2D is shown in Table IV, in which the number of slices in the scan data is fixed to 64 and there are two types of inputs that differ in the way of reading slice images.

The results in Table IV show that both 2D and 3D models can achieve relatively good results on our Clean-CC-CCII dataset, which indicates that the computer-aided COVID-19 diagnosis with state-of-the-art DL techniques would be a promising solution. It shows that DenseNet3D121 is one of the best models among all evaluated models as it achieves the best accuracy, precision, sensitivity, specificity, and F1-score, and MC3_18 obtains the highest AUC score when the slices are read as the greyscale images.

In terms of the accuracy of 3D models, the different number of input channels has different impacts on the different network architectures. However, regarding the AUC metric, almost all 3D models with the greyscale slice images perform better the RGB images. One can see that the ROC curves in Fig. 6 (a) are higher and distributed closer than those in Fig. 6 (b), which indicates that the models trained with greyscale slices are more robust. The main reason is that the original CT slices are greyscale images, and duplicating the greyscale images to RGB images would introduce much repetitive and redundant information, which instead increases the difficulty of model training. Regarding the comparison of 2D models and 3D models, we can see that the overall performance of the 3D models is better than that of the 2D models, which is as expected because the convolutional filters in 3D models can better extract the three-dimensional spatial relationship between the slices of the scan data.

**Fig. 6.**
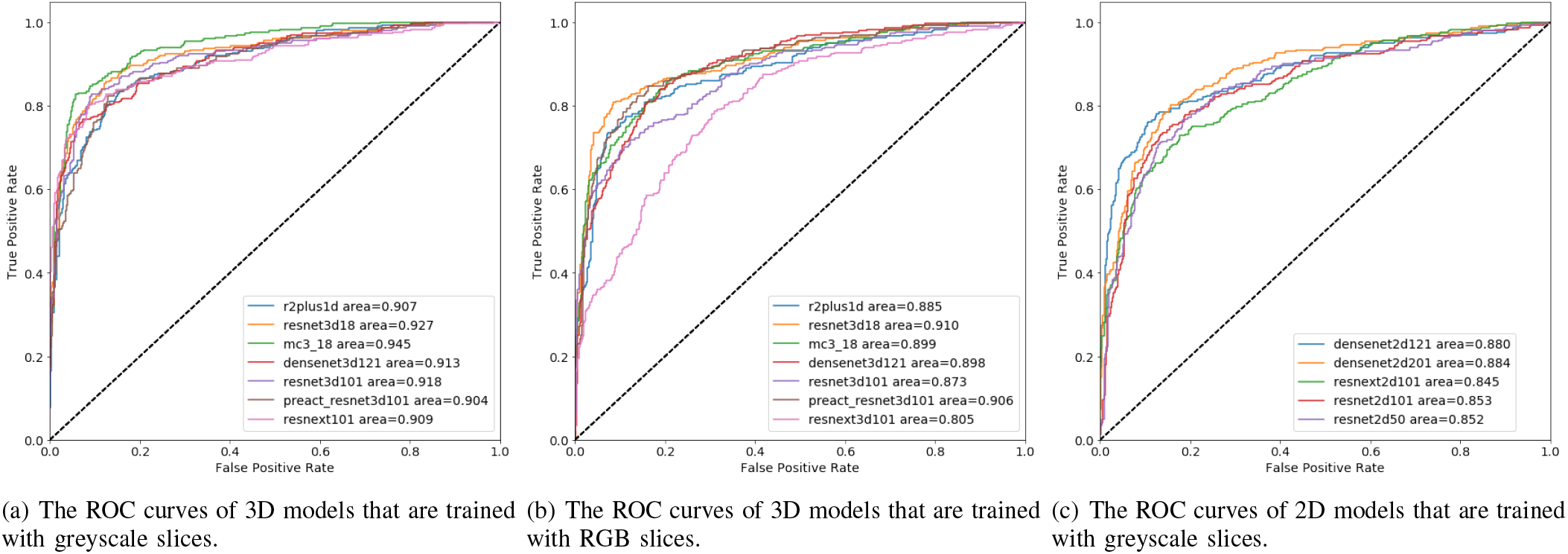
The ROC curves of 3D and 2D models. The overall performance of 3D models is better than 2D models. Besides, the variance between the performance of the models that are trained with greyscale slices is smaller.

We also explore the impact of model depth on model performance as shown in Table V, from which one can see that there is no model that can have an absolute advantages on all metrics. Although no significant correlation can be found between model performance and model depth, the results suggest that a smaller model can also obtain similar or even better results than the larger one.

### C. Results of Exp 2: Comparing the different number of slices

Table VI presents the results of different models trained with the scan data comprising the different number of slices. For MC3 18, DenseNet3D121, and ResNet3D101, we evaluate the cases of 16, 32, 64, 128, and 256 slices, while for ResNeXt3D101, we only test the cases of 16, 32, and 64 slices due to its large size. Fig. 7 (a) plots the relationship between model accuracy and the number of slices. One can see that only the accuracy of ResNet3D101 increases with the number of slices, while other models do not. However, because the distribution of our dataset is imbalanced, higher accuracy does not mean better performance. As Fig. 7 (b) presents, when the number of slices is 64, the AUC of ResNet3D101 is smaller than the other cases. Besides, Fig. 7 (b) also shows that increasing the number of slices does not always improve the performance. Instead, the models trained on a smaller number of slices can also achieve comparable or even better results. For example, both MC3 18 and ResNeXt3D achieves the highest value of AUC when the number of slices is 16.

**TABLE VI.** The performance of models that are trained with scan data comprising of the different number of slices.

| Model | Depth | #Slices | Accuracy (%) | Precision (%) | Sensitivity(Recall) (%) | Specificity (%) | F1-score (%) | AUC |
| --- | --- | --- | --- | --- | --- | --- | --- | --- |
| MC3 | 18 | 16 | 86.78 | **89.00** | **85.76** | **93.55** | **87.35** | **0.936** |
| MC3 | 18 | 32 | 86.16 | 87.11 | 82.78 | 92.54 | 84.89 | 0.913 |
| MC3 | 18 | 64 | 84.75 | 88.34 | 82.78 | 93.35 | 85.47 | 0.890 |
| MC3 | 18 | 128 | **87.50** | 87.76 | 85.43 | 92.74 | 86.58 | 0.920 |
| MC3 | 18 | 256 | 86.12 | 85.91 | 84.77 | 91.53 | 85.33 | 0.908 |
| ResNet3D | 101 | 16 | 85.1 | 86.33 | 79.47 | 92.34 | 82.76 | 0.889 |
| ResNet3D | 101 | 32 | 85.54 | 89.62 | 77.15 | 94.56 | 82.92 | 0.896 |
| ResNet3D | 101 | 64 | 86.00 | 88.19 | 84.11 | 93.15 | 86.1 | 0.873 |
| ResNet3D | 101 | 128 | 87.86 | 89.51 | 84.77 | **93.95** | 87.07 | **0.934** |
| ResNet3D | 101 | 256 | **87.88** | **89.58** | **85.43** | **93.95** | **87.46** | 0.891 |
| ResNeXt3D | 101 | 16 | **86.1** | **88.81** | **84.11** | **93.55** | **86.39** | **0.921** |
| ResNeXt3D | 101 | 32 | 83.94 | 84.83 | 81.46 | 91.13 | 83.11 | 0.909 |
| ResNeXt3D | 101 | 64 | 83.33 | 85.87 | 80.46 | 91.94 | 83.08 | 0.893 |
| DenseNet3D | 121 | 16 | 88.39 | 89.00 | 85.76 | 93.55 | 87.35 | 0.934 |
| DenseNet3D | 121 | 32 | 87.02 | 88.97 | 82.78 | 93.75 | 85.76 | 0.897 |
| DenseNet3D | 121 | 64 | 83.83 | 88.68 | 77.81 | 93.95 | 82.89 | 0.913 |
| DenseNet3D | 121 | 128 | **88.63** | 90.28 | **86.09** | 94.35 | **88.14** | **0.940** |
| DenseNet3D | 121 | 256 | 86.88 | **90.43** | 84.44 | **94.56** | 87.33 | 0.925 |

**Fig. 7.**
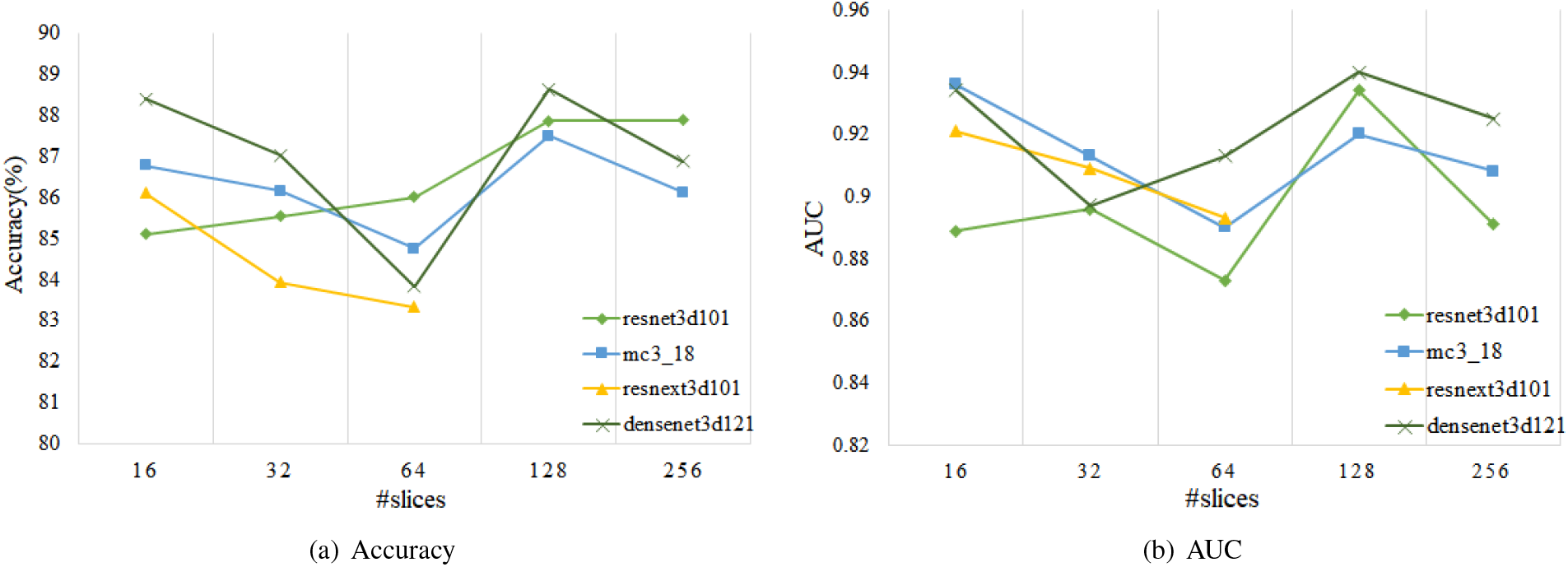
The relationship between model performance and the number of slices. Only the accuracy of ResNet3D101 is positively correlated with the number of slices. When AUC is used as the evaluation metric, there is no obvious relationship between the performance of the model and the number of slices.

### D. Results of Exp 3: Training with mixup

Table VIII presents the model performance before and after using the mixup strategy. One can see that mixup can significantly enhance model performance. After applying mixup, the accuracy of R2Plus1D18, MC3 18, DenseNet3D121, and ResNet3D101 is improved by 3.00%, 1.44%, 4.27%, and 2.64%, respectively.

**TABLE VII.**
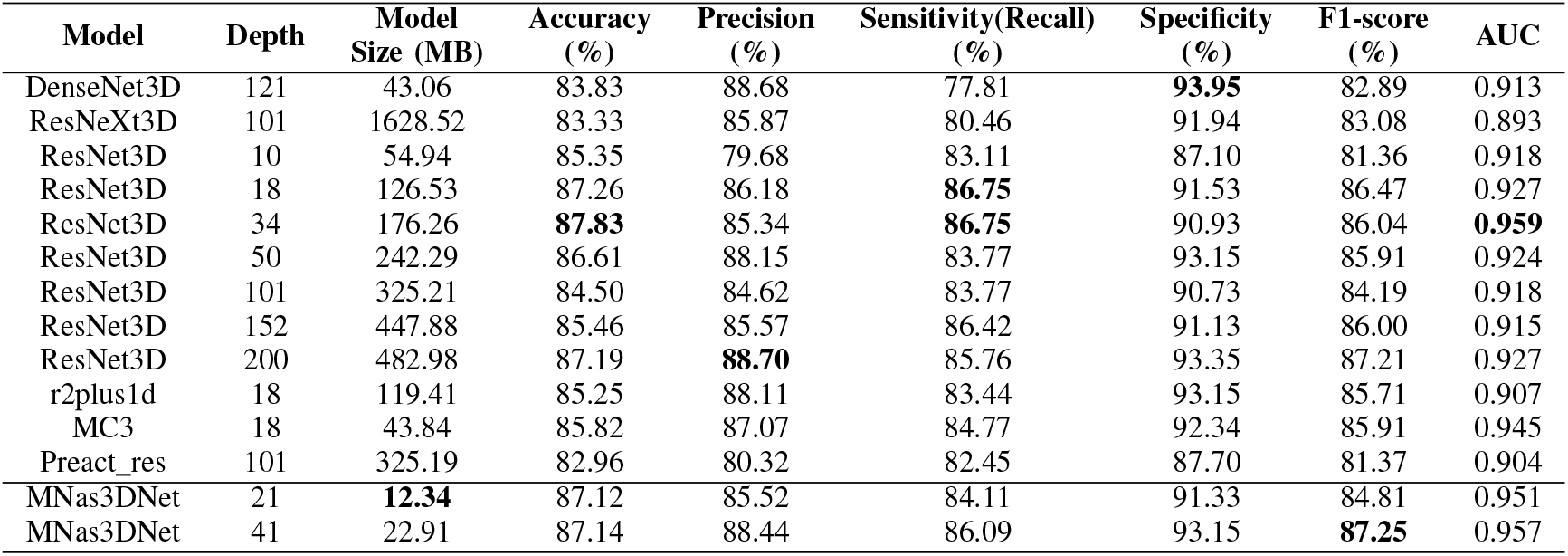
The performance comparison between the baseline models and the models searched by NAS. For all models, the input scan images are composed of 64 greyscale slice images.

| Model | Depth | Model Size (MB) | Accuracy (%) | Precision (%) | Sensitivity(Recall) (%) | Specificity (%) | F1-score (%) | AUC |
| --- | --- | --- | --- | --- | --- | --- | --- | --- |
| DenseNet3D | 121 | 43.06 | 83.83 | 88.68 | 77.81 | **93.95** | 82.89 | 0.913 |
| ResNeXt3D | 101 | 1628.52 | 83.33 | 85.87 | 80.46 | 91.94 | 83.08 | 0.893 |
| ResNet3D | 10 | 54.94 | 85.35 | 79.68 | 83.11 | 87.10 | 81.36 | 0.918 |
| ResNet3D | 18 | 126.53 | 87.26 | 86.18 | **86.75** | 91.53 | 86.47 | 0.927 |
| ResNet3D | 34 | 176.26 | **87.83** | 85.34 | **86.75** | 90.93 | 86.04 | **0.959** |
| ResNet3D | 50 | 242.29 | 86.61 | 88.15 | 83.77 | 93.15 | 85.91 | 0.924 |
| ResNet3D | 101 | 325.21 | 84.50 | 84.62 | 83.77 | 90.73 | 84.19 | 0.918 |
| ResNet3D | 152 | 447.88 | 85.46 | 85.57 | 86.42 | 91.13 | 86.00 | 0.915 |
| ResNet3D | 200 | 482.98 | 87.19 | **88.70** | 85.76 | 93.35 | 87.21 | 0.927 |
| r2plus1d | 18 | 119.41 | 85.25 | 88.11 | 83.44 | 93.15 | 85.71 | 0.907 |
| MC3 | 18 | 43.84 | 85.82 | 87.07 | 84.77 | 92.34 | 85.91 | 0.945 |
| Preact_res | 101 | 325.19 | 82.96 | 80.32 | 82.45 | 87.70 | 81.37 | 0.904 |
| MNas3DNet | 21 | **12.34** | 87.12 | 85.52 | 84.11 | 91.33 | 84.81 | 0.951 |
| MNas3DNet | 41 | 22.91 | 87.14 | 88.44 | 86.09 | 93.15 | **87.25** | 0.957 |

**TABLE VIII.** The model accuracy before and after using the mixup strategy. The number of slices is fixed to 64 for all models.

| Model | Accuracy(%) |  | Improvement (%) |
| --- | --- | --- | --- |
|  | before mixup | after mixup |  |
| R2Plus1D18 | 85.25 | 88.25 | +3.00 |
| MC3_18 | 85.82 | 87.26 | +1.44 |
| DenseNet3D121 | 83.83 | 88.10 | +4.27 |
| ResNet3D101 | 84.50 | 87.14 | +2.64 |

A possible explanation for this result might be that the original training data can be regarded as a pile of scattered points distributed in high-dimensional space, and a large number of new data points between the original data points are created by the mixup method. In this way, the original dataset is expanded to some extent, and the data distribution becomes smoother, which regularizes the model training and improves the model performance.

### E. Results of automated model design for COVID-19 detection

We implement two types of NAS experiments. One is to search for 21-layer networks, taking 3.7 hours, while the other searching for 41-layer networks took 5 hours. Table VII presents the performance comparison between the baseline 3D models and our searched 3D models by NAS, namely MNas3DNet. To have a fair comparison, for all models, the input scan images are composed of 64 greyscale slice images. Compared to the baseline 3D models, the sizes of our searched models are much smaller, where MNas3DNet21 and MNas3DNet41 are 12.34 and 22.91 MB, respectively. At the same time, both models achieve the SOTA performance. Specifically, MNas3DNet41 achieves an accuracy of 87.14%, F1-score of 87.25%, and AUC of 0.957, which are on par with the best models designed by AI experts. The strong empirical results prove the effectiveness of random search strategy, and demonstrate that NAS is a promising research direction for designing neural networks of detecting COVID-19.

## V. Conclusion and future work

In this paper, we aim to benchmark DL models and use AutoML techniques to design DL models for COVID-19 detection using chest CT scans. Our experimental results show that DL models are promising solutions, and 3D models outperform 2D models. We find that the model performance does not absolutely improve with the increase of model depth or the number of slices. In other words, a smaller model trained on less number of slices can also achieve comparable or even better results. Besides, we demonstrate that mixup data augmentation can effectively improve model performance. Last but not least, we design an automated deep learning methodology to generate a lightweight deep learning model, which achieves comparable results to the models designed by AI experts.

We have several directions for future work on the agenda as follows. First, most of the data in our dataset are from China, thus we plan to collect more data from other countries to further improve the accuracy of COVID-19 detection. Second, we will try to apply semantic segmentation technology to our dataset, so as to help doctors diagnose more effectively. Last, we will try other SOTA NAS methods to explore more types of deep learning models.

## Data Availability

Our data is obtained from a publicly available dataset, i.e., http://ncov-ai.big.ac.cn/download?lang=en, which is under a Creative Commons Attribution 3.0 China Mainland License.

https://github.com/HKBU-HPML/HKBU_HPML_COVID-19

http://ncov-ai.big.ac.cn/download?lang=en

## Appendix

### A. Slice Sampling Strategies

#### 1) Random Sampling Strategy

The random sampling strategy is applied to the training set. In this way, each scan data will be composed of different slices, which can be regarded as data augmentation to improve the model robustness and avoid overfitting.

##### Algorithm 1

Random slice sampling algorithm.

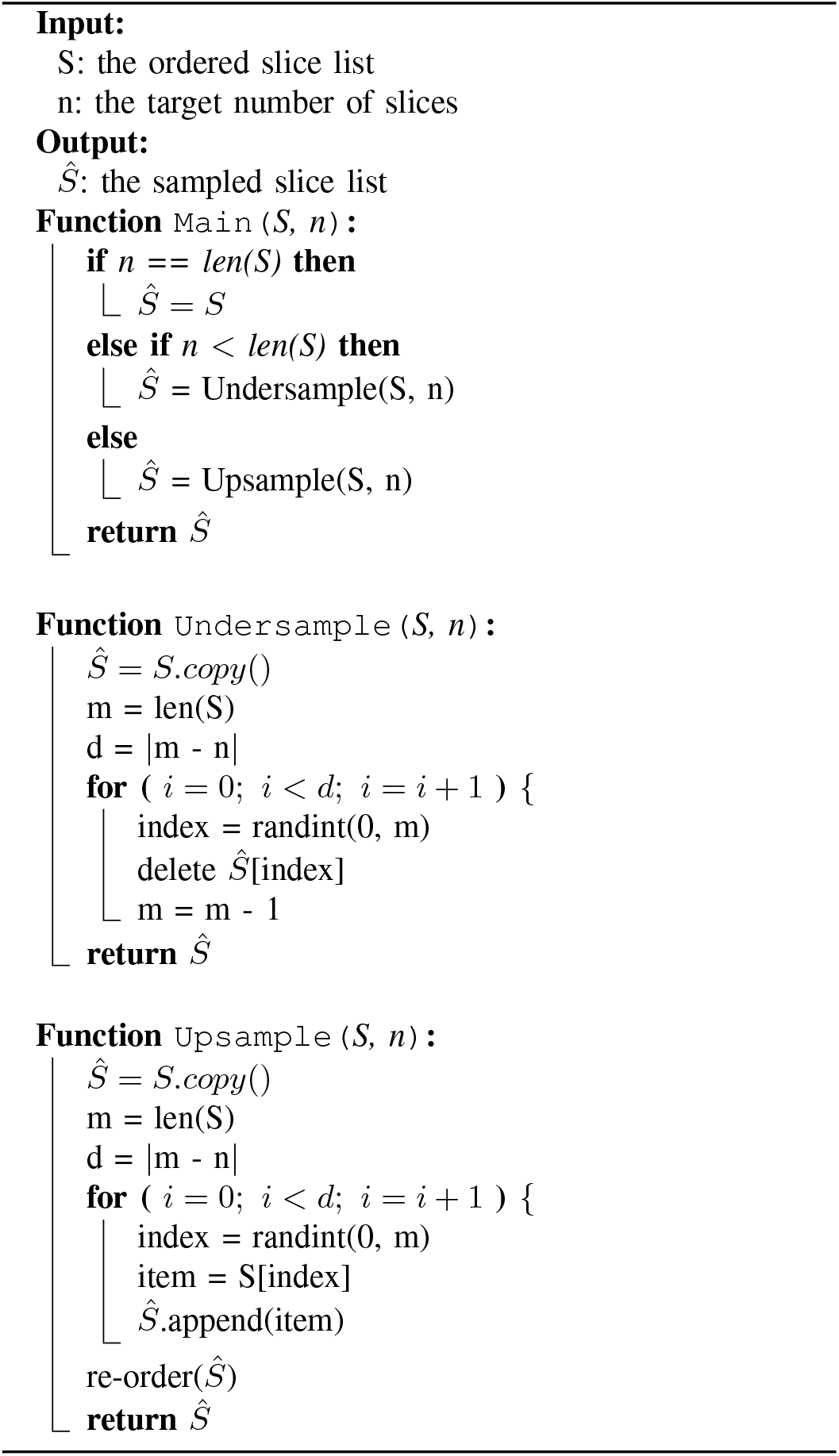

#### B. Symmetrical Sampling Strategy

The symmetrical sampling strategy is applied to the test set. This avoids the randomness of the test results, and also makes a fair performance comparison between different models.

##### Algorithm 2

Symmetrical slice sampling algorithm.

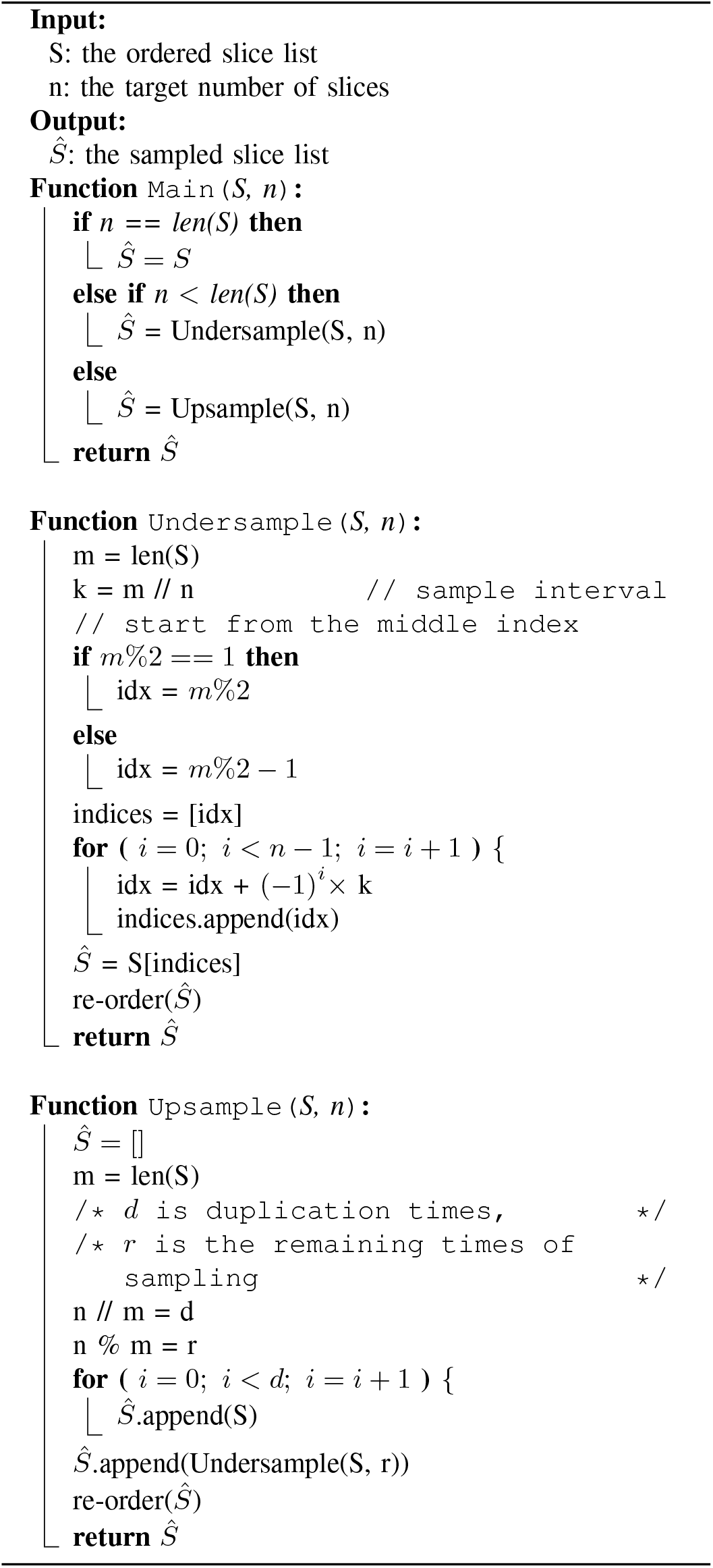

## Footnotes

1 We found some errors and noises in the dataset, and hence built our own version.

2 https://github.com/booz-allen-hamilton/DSB3Tutorial

3 https://github.com/microsoft/nni

## Notes

### Competing Interest Statement

The authors have declared no competing interest.

### Clinical Protocols

https://github.com/HKBU-HPML/HKBU_HPML_COVID-19

### Funding Statement

nil

### Summary of Updates

update the URL of the source code

## References

[1] W. H. Organization et al., “Naming the coronavirus disease (covid-19) and the virus that causes it,” World Health Organization. https://www.who.int/emergencies/diseases/novel-coronavirus-2019/technical-guidance/naming-the-coronavirus-disease-(covid-2019)-and-the-virus-that-causes-it, 2020.

[2] W. H. Organization, “Q&a on coronaviruses (covid-19),” World Health Organization, 2020.

[3] A. Chin, J. Chu, M. Perera, K. Hui, H.-L. Yen, M. Chan, M. Peiris, and L. Poon, “Stability of SARS-CoV-2 in different environmental conditions,” medRxiv, 2020.

[4] W. H. Organization et al., “Coronavirus disease (covid-2019) situation reports,” Accessd 3 June 2020. [Online]. Available: https://www.who.int/emergencies/diseases/novel-coronavirus-2019/situation-reports

[5] T. Ai, Z. Yang, H. Hou, C. Zhan, C. Chen, W. Lv, Q. Tao, Z. Sun, and L. Xia, “Correlation of chest ct and rt-pcr testing in coronavirus disease 2019 (covid-19) in china: a report of 1014 cases,” Radiology, p. 200642, 2020.

[6] K. Zhang, X. Liu, J. Shen, Z. Li, Y. Sang, X. Wu, Y. Zha, W. Liang, C. Wang, K. Wang et al., “Clinically applicable AI system for accurate diagnosis, quantitative measurements, and prognosis of covid-19 pneumonia using computed tomography,” Cell, 2020.

[7] J. Zhang, Y. Xie, Y. Li, C. Shen, and Y. Xia, “COVID-19 Screening on Chest X-ray Images Using Deep Learning based Anomaly Detection,” 2020. [Online]. Available: http://arxiv.org/abs/2003.12338

[8] B. Ghoshal and A. Tucker, “Estimating Uncertainty and Interpretability in Deep Learning for Coronavirus (COVID-19) Detection,” pp. 1–14, 2020. [Online]. Available: http://arxiv.org/abs/2003.10769

[9] A. Narin, C. Kaya, and Z. Pamuk, “Automatic Detection of Coronavirus Disease (COVID-19) Using X-ray Images and Deep Convolutional Neural Networks,” 2020. [Online]. Available: http://arxiv.org/abs/2003.10849

[10] D. Singh, V. Kumar Vaishali, and M. Kaur, “Classification of COVID-19 patients from chest CT images using multi-objective differential evolution-based convolutional neural networks,” European journal of clinical microbiology & infectious diseases : official publication of European Society of Clinical Microbiology, 2020.

[11] A. A. Ardakani, A. R. Kanafi, U. R. Acharya, N. Khadem, and A. Mohammadi, “Application of deep learning technique to manage COVID-19 in routine clinical practice using CT images: Results of 10 convolutional neural networks,” Computers in Biology and Medicine, vol. 121, no. March, p. 103795, 2020. [Online]. Available: http://www.sciencedirect.com/science/article/pii/S0010482520301645

[12] M. Z. Alom, M. M. S. Rahman, M. S. Nasrin, T. M. Taha, and V. K. Asari, “Covid MTNet: Covid-19 detection with multi-task deep learning approaches,” 2020.

[13] X. He, X. Yang, S. Zhang, J. Zhao, Y. Zhang, E. Xing, and P. Xie, “Sample-Efficient Deep Learning for COVID-19 Diagnosis Based on CT Scans,” medRxiv, vol. XX, no. Xx, p. 2020.04.13.20063941, 2020.

[14] A. Mobiny, P. A. Cicalese, S. Zare, P. Yuan, M. Abavisani, C. C. Wu, J. Ahuja, P. M. de Groot, and H. Van Nguyen, “Radiologist-Level COVID-19 Detection Using CT Scans with Detail-Oriented Capsule Networks,” 2020. [Online]. Available: http://arxiv.org/abs/2004.07407

[15] C. Zheng, X. Deng, Q. Fu, Q. Zhou, J. Feng, H. Ma, W. Liu, and X. Wang, “Deep Learning-based Detection for COVID-19 from Chest CT using Weak Label,” medRxiv, p. 2020.03.12.20027185, 2020. [Online]. Available: http://medrxiv.org/content/early/2020/03/17/2020.03.12.20027185

[16] L. Li, L. Qin, Z. Xu, Y. Yin, X. Wang, B. Kong, J. Bai, Y. Lu, Z. Fang, Q. Song et al., “Artificial intelligence distinguishes covid-19 from community acquired pneumonia on chest ct,” Radiology, p. 200905, 2020.

[17] K. Hara, H. Kataoka, and Y. Satoh, “Can spatiotemporal 3D CNNs retrace the history of 2D CNNs and ImageNet?” in Proceedings of the IEEE conference on Computer Vision and Pattern Recognition, 2018, pp. 6546–6555.

[18] D. Tran, H. Wang, L. Torresani, J. Ray, Y. LeCun, and M. Paluri, “A closer look at spatiotemporal convolutions for action recognition,” 2017.

[19] G. Huang, Z. Liu, L. Van Der Maaten, and K. Q. Weinberger, “Densely connected convolutional networks,” in Proceedings of the IEEE conference on computer vision and pattern recognition, 2017, pp. 4700–4708.

[20] K. He, X. Zhang, S. Ren, and J. Sun, “Deep residual learning for image recognition,” in Proceedings of the IEEE conference on computer vision and pattern recognition, 2016, pp. 770–778.

[21] S. Xie, R. Girshick, P. Dollár, Z. Tu, and K. He, “Aggregated residual transformations for deep neural networks,” in Proceedings of the IEEE conference on computer vision and pattern recognition, 2017, pp. 1492– 1500.

[22] X. He, K. Zhao, and X. Chu, “Automl: A survey of the state-of-the-art,” arXiv preprint 1908.00709, 2019.

[23] F. Hutter, L. Kotthoff, and J. Vanschoren, “Automated machine learning: Methods, systems, challenges,” Automated Machine Learning, 2019.

[24] H. Zhang, M. Cisse, Y. N. Dauphin, and D. Lopez-Paz, “mixup: Beyond empirical risk minimization,” 2017.

[25] G. Litjens, T. Kooi, B. E. Bejnordi, A. A. A. Setio, F. Ciompi, M. Ghafoorian, J. A. van der Laak, B. van Ginneken, and C. I. Sánchez, “A survey on deep learning in medical image analysis,” Medical Image Analysis, vol. 42, no. 1995, pp. 60–88, 2017.

[26] J. P. Cohen, P. Morrison, and L. Dao, “Covid-19 image data collection,” 2003.11597, 2020. [Online]. Available: https://github.com/ieee8023/covid-chestxray-dataset

[27] J. Zhao, X. He, X. Yang, Y. Zhang, S. Zhang, and P. Xie, “Covid-ct-dataset: A ct scan dataset about covid-19,” 2020.

[28] M. Jun, G. Cheng, W. Yixin, A. Xingle, G. Jiantao, Y. Ziqi, Z. Minqing, L. Xin, D. Xueyuan, C. Shucheng, W. Hao, M. Sen, Y. Xiaoyu, N. Ziwei, L. Chen, T. Lu, Z. Yuntao, Z. Qiongjie, D. Guoqiang, and H. Jian, “COVID-19 CT Lung and Infection Segmentation Dataset,” Apr. 2020. [Online]. Available: https://doi.org/10.5281/zenodo.3757476

[29] T. Elsken, J. H. Metzen, and F. Hutter, “Neural architecture search: A survey,” arXiv preprint 1808.05377, 2018.

[30] L. Faes, S. K. Wagner, D. J. Fu, X. Liu, E. Korot, J. R. Ledsam, T. Back, R. Chopra, N. Pontikos, C. Kern, G. Moraes, M. K. Schmid, D. Sim, K. Balaskas, L. M. Bachmann, A. K. Denniston, and P. A. Keane, “Automated deep learning design for medical image classification by health-care professionals with no coding experience: a feasibility study,” The Lancet Digital Health, vol. 1, no. 5, pp. e232–e242, 2019. [Online]. Available: http://dx.doi.org/10.1016/S2589-7500(19)30108-6

[31] A. G. Howard, M. Zhu, B. Chen, D. Kalenichenko, W. Wang, T. Weyand, M. Andreetto, and H. Adam, “Mobilenets: Efficient convolutional neural networks for mobile vision applications,” 2017.

[32] M. Sandler, A. Howard, M. Zhu, A. Zhmoginov, and L.-C. Chen, “Mobilenetv2: Inverted residuals and linear bottlenecks,” in Proceedings of the IEEE conference on computer vision and pattern recognition, 2018, pp. 4510–4520.

[33] M. Tan, B. Chen, R. Pang, V. Vasudevan, M. Sandler, A. Howard, and Q. V. Le, “Mnasnet: Platform-aware neural architecture search for mobile,” in Proceedings of the IEEE Conference on Computer Vision and Pattern Recognition, 2019, pp. 2820–2828.

[34] B. Wu, X. Dai, P. Zhang, Y. Wang, F. Sun, Y. Wu, Y. Tian, P. Vajda, Y. Jia, and K. Keutzer, “Fbnet: Hardware-aware efficient convnet design via differentiable neural architecture search,” in Proceedings of the IEEE Conference on Computer Vision and Pattern Recognition, 2019, pp. 10 734–10 742.

[35] L. Li and A. Talwalkar, “Random search and reproducibility for neural architecture search,” arXiv preprint 1902.07638, 2019.

[36] C. Sciuto, K. Yu, M. Jaggi, C. Musat, and M. Salzmann, “Evaluating the search phase of neural architecture search,” arXiv preprint 1902.08142, 2019.

[37] D. P. Kingma and J. Ba, “Adam: A method for stochastic optimization,” 2014.

[38] I. Loshchilov and F. Hutter, “SGDR: Stochastic gradient descent with warm restarts,” International Conference on Learning Representations (ICLR), 2017.

